# Parents’ experiences and information needs when seeking emergency department care for their medically complex child: a mixed methods systematic review protocol

**DOI:** 10.1101/2025.07.09.25331190

**Authors:** Sarah A Elliott, Lisa Hartling, Samina Ali, Danielle Lysak, Deborah Olmstead, Lisa Knisley, Liz Dennett, Hannah M Brooks, Shannon D Scott

**Affiliations:** Department of Pediatrics, Faculty of Medicine and Dentistry, University of Alberta, Edmonton, Alberta, Canada; Faculty of Nursing, University of Alberta, Edmonton, Alberta, Canada; Geoffrey and Robyn Sperber Health Sciences Library, University of Alberta, Edmonton, Alberta, Canada; Stollery Children’s Hospital, Edmonton, Alberta, Canada; College of Nursing, Rady Faculty of Health Sciences, University of Manitoba, Winnipeg, Manitoba, Canada

**Keywords:** experiences, information needs, systematic review, mixed methods, pediatrics, medical complexity

## Abstract

**Objective:** To provide a comprehensive understanding of parents’ self-reported experiences and/or information needs related to the management of their child with medical complexity in the emergency department.

**Background:** Children with medical complexity frequently rely on emergency department services to address sudden changes in health status. While some primary research explores family experiences in caring for these children, there has been no comprehensive synthesis of the available evidence.

**Inclusion Criteria:** Primary studies of any design will be included (quantitative, qualitative, mixed-method, and multi-method) if they are conducted in outpatient care settings, ambulatory care, or primary care settings (emergency departments, urgent care centres, community clinics) with parents, family caregivers, or legal guardians of children (0-18 years) with medical complexity. Studies must present parents’ self-reported experiences and/or information needs related to the management of their child with medical complexity.

**Methods:** A health research librarian will develop and implement search strategies in the following electronic databases: Ovid MEDLINE, Ovid Embase, Cochrane Library, Ovid PsycINFO, and CINAHL Plus with Full Text via EBSCOhost to identify relevant evidence. Two reviewers will independently screen and categorize identified records based on titles and abstracts (Stage 1). The full text of relevant records will then be screened independently by two reviewers (Stage 2). Quality assessments and data extraction of the included records will be done in duplicate using standardized forms. Disagreements will be resolved through third-party adjudication. The Mixed-Methods Appraisal Tool version 2018 (MMAT) will be used to assess the methodological quality of included studies. Data analysis will follow the Joanna Briggs Institute (JBI) convergent integrated approach, whereby quantitative and qualitative data will be synthesized simultaneously.

**Summary:** Findings from this review may be utilised by healthcare providers and policy makers to enhance parents’ experiences and support the care of children with medical complexity in emergency settings. These findings may also inform the development of knowledge translation resources (e.g., videos, infographics) for parents, as well as educational tools or interventions for healthcare providers working in emergency departments.

**Review Registration:** The a priori protocol for this mixed methods systematic review was registered with PROSPERO on April 26, 2024 (PROSPERO CRD42024534701).

## Introduction

Children with medical complexity share four defining characteristics: they have one or more complex, chronic multisystem conditions; they experience functional limitations that require reliance on technology; frequent utilization of healthcare services across multiple providers; and high overall healthcare needs [1]. Due to the multisystem nature of their conditions, children with medical complexity are at increased risk of complications, frequently requiring care in the emergency department (ED) [2].

Given that ED visits are often triggered by rapid clinical changes, the experience is overwhelming for all involved. Challenges in the ED are further exacerbated by the unique complexities of each child with medical complexity, their functional abilities, and their acute needs. During ED visits, parents must relay extensive health histories while simultaneously managing their child’s acute needs, adding to the stress of urgent care encounters [3–5]. However, there remains limited understanding both in the literature and in practice of how parents of children with medical complexity navigate these visits. To bridge this gap, it is crucial to examine their experiences and information needs in ED settings. Investigating these perspectives can identify key areas for improving communication, care management, and family support.

### Review questions

1. What are parents’ self-reported experiences relating to the management of their child with complex medical needs in the ED?

2. What are parents’ self-reported information needs related to the management of their child with complex medical needs in the ED?

### Inclusion Criteria

#### Participants

This review will consider studies including parents, familiar caregivers, or legal guardians of children (0-18 years of age) with medical complexity. In order for children to be considered medically complex there are four defining characteristics: 1) one or more complex chronic multisystem conditions; 2) functional limitations that cause reliance on technology; 3) high health care utilization with multiple providers and 4) high health care service needs.

#### Phenomena of Interest

This review will consider studies that explore parents’ self-reported experiences and/or information needs related to the management of their child with medical complexity. For this review, we define experiences as how parents feel (e.g., overwhelmed, confident) and act (e.g., supportive, withdrawn) before, during and after their child attends the ED. Experiences are deeply subjective in nature and encompass a wide range of physical, psychological, and/or emotional events or circumstances that are unique to the individual, and may be influenced by social cognitive factors, such as attitudes and beliefs, or demographic factors, such as culture [6, 7]. Thus, they should be measured through a careful and comprehensive approach [7]. Information needs are defined as the type, content or topic, quantity, frequency, and mode of delivery of information that parents require or desire to receive (parents’ preferences). Information needs are likely to be influenced by parents’ health-care related experiences and their existing state of knowledge, which in turn may influence their decision-making. Thus, parent’s information needs may include desire for information to address any knowledge gap that may exist.

#### Context

This review will consider studies conducted in outpatient care settings (e.g., ambulatory and primary care including emergency departments, urgent care centers, and community clinics), regardless of geographic location. We will not limit study inclusion to any defined sub-population of parents of children with medical complexity.

#### Types of studies

This review will consider primary quantitative, qualitative, mixed methods and multi method studies. We will exclude intervention studies (e.g., educational interventions, behavioral modification interventions) that aimed to influence parents’ experiences or information needs in relation to their experiences in the emergency department. However, if the study reports baseline data related to parents’ experiences and/or information needs, we will include that data only.

## Methods

The proposed systematic review will be conducted in accordance with the JBI methodology for mixed methods systematic reviews for a convergent integrated approach [8] and follows methods used in our previously published mixed methods reviews [9–11]. By employing a convergent integrated approach [8] combining qualitative and quantitative evidence within a single review, the resultant synthesis will enhance our insight into the experiences and information needs of parents caring for a child with medical complexity requiring ED care [8, 12, 13]. We will report the findings of the review in line with the Preferred Reporting Items for Systematic Reviews and Meta-Analyses (PRISMA) guidelines [14].

### Search Strategy

A health sciences librarian (Dennett) will develop and refine the search strategy in consultation with methodologists (Elliott, Hartling, Scott) and content experts (Ali, Lysak, Olmstead). Initial scoping will be conducted using MEDLINE and Embase, to ensure appropriate terms are included (e.g., medical complexity, chronic disease, cancer, child, parent and caregivers, information seeking behaviour, experience, perception). The search strategy will be peer reviewed using the Peer Review of Electronic Search Strategies (PRESS) checklist [15] by a second librarian with systematic review experience. Once finalised, the search strategy will be translated for use in each electronic database to be searched, including: Ovid MEDLINE, Ovid Embase, Cochrane Library, Ovid PsycINFO, and CINAHL Plus with Full Text via EBSCO host from inception to May 2024. This search will be restricted to studies published in English, and although this is not expected to limit results [16], it may affect the generalizability of findings to practice settings. As health systems vary substantially by location and funding mechanism, we expect there may be limitations to the generalizability of results.

Relevant grey literature will be located by searching ProQuest Dissertations and Theses Global. Meeting abstracts from the past two years will be searched for in the Conference Proceedings Citation Index (Clarivate Analytics). We will flag relevant reviews/overviews to scan their reference lists and identify any additional studies relevant to our review that were not captured in our search.

### Study Selection

Following the search, all identified citations will be imported into Covidence and duplicates removed. Following a pilot test to ensure adequate understanding of the inclusion/exclusion criteria, two independent reviewers will screen and categorize identified records based on titles and abstracts (Stage 1). Each record will be classified as “Include/Unsure” or “Exclude”, guided by the eligibility criteria. Next, full text articles of all records classified as “Include/Unsure” will be retrieved and reviewed for inclusion by two independent reviewers (stage 2). Discrepancies will be resolved through third-party adjudication. If full text is unavailable to be retrieved, we will contact the corresponding author twice and request access to the full text. If they do not respond, we will exclude the abstract. A PRISMA flow diagram [17] will be used to summarize search results and selection processes.

### Assessment of methodological quality

The Mixed-Methods Appraisal Tool version 2018 (MMAT) [18] will be used to assess the methodological quality of included studies. Two reviewers will independently assess the quality of each included study and provide justification for their decisions. A pilot test on approximately five records will be conducted to ensure consistency before appraisal begins. Authors of papers will be contacted to request missing or additional data for clarification, where required. Any disagreements that arise between the reviewers will be resolved through discussion or with a third reviewer. All studies will be included, regardless of quality assessment score, to ensure a comprehensive representation of parent reported experiences and information needs related to their child’s management. The results of critical appraisal will be reported in narrative format and in a table.

### Data extraction

Data extraction forms have previously been developed and will be refined following pilot testing using a sample of 10 full text articles. Data extraction will include: study details and characteristics (e.g., authors/organizations, publication year, country, study design), participant characteristics including intersectionality data on social identities (e.g., age, race, gender, parental role) and clinical conditions (e.g., cerebral palsy, difficulty swallowing with a urological condition; genetic syndrome with associated congenital heart defect), and reported outcomes related to parents’ experiences and information needs. To conceptualize experiences and information needs, we will adopt previously used definitions that our research team has employed in similar knowledge synthesis research of other pediatric health conditions [9–11, 19–22]. Data will be extracted by one reviewer and verified by a second reviewer. Disagreements will be discussed and resolved by consensus.

Relevant quantitative data will be extracted directly into Covidence. Relevant qualitative data including participant quotes or authors’ reported data in ‘findings’ or ‘results’ sections of the included studies will be extracted verbatim into NVivo data management software by one reviewer and verified by a second reviewer.

### Data Transformation

Quantitative results will be converted into “qualitized” data by transforming quantitative data into textual descriptions or narrative interpretation. One reviewer will complete this step, and a second reviewer will confirm the conversions.

### Data Synthesis and integration

Quantitative and qualitative data will be synthesized simultaneously using a convergent integrated approach [8, 13]. Using NVivo data management software, one reviewer will code the qualitative data inductively using thematic analysis by applying one or more codes to each line of text according to its meaning and content. All preliminary codes will be verified by a second reviewer to reduce the risk of interpretive bias. Codes will then be categorized into themes and sub-themes. This step will be repeated until no new themes emerge from the data. Emergent themes and sub-themes will be finalized through discussion with the research team.

To create a set of integrated findings, our team will utilize a methodological approach similar to one we have previously employed to answer similar research questions [10, 11, 21]. We will pool the “qualitized” and qualitative data that have similar meanings and contents, where possible. If pooling is not possible, sub-themes will be generated based on one data format while the other data format will be used to complement findings. The qualitative data and the data that has been “qualitized” will be constantly compared, coded, and categorized into themes throughout the integration process, until no new themes emerge. An independent reviewer experienced in both quantitative and qualitative methods will review, verify and discuss the final integrated themes and sub-themes with the research team.

### Analysis of subgroups or subsets

When available, we will record participant features from included studies (as outlined in data extraction tables) to help identify communities or groups of children that may benefit from additional research efforts in the future.

## Summary

Children with medical complexity live across a spectrum of complexity, functional abilities, communication abilities, and healthcare needs. As such, the impact on families is widespread and varied [3]. As with all studies involving vulnerable populations, we expect that experiences of parents of children with medical complexity will vary widely given the intersectional characteristics of individuals and their families. We expect findings to synthesize available experiences and information needs of families across the continuum of intersections and complexities.

Furthermore, we expect findings will offer significant insights into systemic and individual (both parent and healthcare provider) aspects of care that can be improved. Through the identification and synthesis of findings, we hope to inform future knowledge translation efforts aimed at improving patient-centered care and supporting positive experiences in the ED for parents of children with medical complexity.

## Data Availability

Data extracted from included studies is available upon reasonable request to the corresponding author.

## Appendix I Search strategy

Ovid MEDLINE(R) ALL <1946 to [search date]>

Search conducted: [add date]

Records retrieved: [add number]

1. chronic disease/ or critical illness/ or rare diseases/ or Palliative Care/ or Palliative Medicine/ or exp Spinal Dysraphism/ or exp Anemia, Sickle Cell/ or exp neoplasms/ or Cerebral Palsy/ or exp abnormalities, multiple/ or exp 22q11 deletion syndrome/ or Transplant Recipients/ or exp Heart Defects, Congenital/ or exp Infant, Premature/ or muscular atrophy, spinal/ or “spinal muscular atrophies of childhood”/
2. (“medical complexity” or “complex medical” or “medically complex” or “medically fragile” or “medical fragility” or “complex health” or “complex conditions” or “complex diseases” or “complex chronic” or “complex care” or “multiple comorbidities” or ((complex or special) adj4 needs) or technology-dependen* or life-threatening or life-limiting or “complex pediatric” or “complex paediatric” or palliative).mp.
3. (“chronic disease*” or “chronic* ill*” or “chronic condition*” or “chronic disorder*” or “rare disease*” or “rare disorder*” or “rare conditions” or ((genetic or hereditary or chromosome) adj2 (disease* or disorder*)) or “rare genetic condition*” or (degenerative adj3 (disease* or disab* or ill* or condition)) or “deletion syndrome*” or “chromosome deletion” or tracheostom* or “artificial airway*” or “invasive ventilation*” or “ventilator dependent” or “spina bifida” or “sickle cell” or “motor neuron disease*” or “spinal muscular atrophy” or “cerebral palsy” or “global developmental delay*” or transplant recipient* or ((await* or list or candidat*) adj3 transplant*) or “congenital heart disease*” or “complex heart disease*” or “premature infant*” or “preterm infant*” or “premature babies” or “premature baby” or preemies or micropreemies or “premature birth*” or “extreme* prematur*”).mp.
4. (cancer* or neoplas* or tumour* or tumor* or leukemia* or sarcoma* or rhabdomyosarcoma* or osteosarcoma* or melanoma* or myeloma* or lymphoma* or carcinoma* or astrocytoma* or medulloblastoma* or neuroblastoma* or blastoma* or retinoblastoma* or esthesioneuroblastoma* or histiocyto* or craniopharyngioma* or glioma* or ependymoma* or mesothelioma* or malignan* or oncolog* or papillomatos* or paraganglioma* or pheochromocytoma).mp.
5. or/1-4
6. adolescent/ or exp child/ or exp infant/
7. ((child* or teen* or adolesc* or preteen* or youth or youths or toddler* or infant* or baby or babies or newborn* or neonat* or preschool* or pre-school* or pediatric* or paediatric* or kids or boys or girls or son or sons or daughter*) not adult-children).mp.
8. 6 or 7
9. exp Parents/ or Family/ or Caregivers/
10. (caregiver* or care-giver* or guardian* or parent or parents or parental or mother* or maternal or father* or paternal or moms or dads or family or families or grandparent*).mp.
11. 9 or 10
12. ((caregiver* or care-giver* or guardian* or parent or parents or parental or mother or mothers or maternal or father or fathers or paternal or moms or dads or family or families or grandparent* or child* or teen* or adolesc* or preteen* or youth* or toddler* or infant* or baby or babies or newborn* or neonat* or preschool* or pre-school* or kids or boys or girls or son or sons or daughter*) adj6 (experience* or perception* or perceive* or perspective* or opinion* or attitude* or belief* or voice or self-report* or self-confiden* or self-efficac* or narrati* or expectation* or impression* or views or stories or reflections or preferences or complian* or comply* or noncomplian* or adhere* or nonadhere* or awareness or satisfied or satisfaction or faciltitat* or challenges or barrier* or difficulties or difficulty or obstacle* or hurdle* or barricade* or hindrance* or obstruct* or disparit* or inequi* or inequal* or impede* or impediment* or interview* or focus-group* or questionnaire* or survey* or anxiety or anxious* or concern* or unsure or uncomfortable or comfortable or happy or unhappy or complain* or stress* or worry or worried or fear* or distress* or thoughts or feeling* or fright* or helpless* or sleepless* or emotion* or misconception* or misperce* or misunderstand* or myth* or understand* or understood or unaware* or preparedness or involvement)).mp.
13. ((need or needs or seek* or preference* or request* or support or supports) adj6 (information or knowledge or learn* or comprehension or comprehend* or understand* or train* or educat*)).mp.
14. ((know* or learn* or comprehen* or information or guidance or instruct*) adj4 (more or lack or lacking or missing or better or deficit* or deficient* or sufficient or insufficient or interested or interest or interesting or barrier* or challeng* or difficulty or difficulties or obstacle* or discourage* or disparit* or enhance or facilitat* or gap or gaps or hindrance or hurdle or imped* or inequit* or inequalit* or increas*)).mp.
15. consumer health information/ or patient education as topic/ or needs assessment/
16. Caregivers/ed, px or Family/ed, px or Parents/ed, px
17. Attitude to health/ or Health Knowledge, Attitudes, Practice/ or health literacy/ or exp patient satisfaction/
18. Information Seeking Behavior/
19. (literacy or literate or illitera*).mp.
20. ((caregiver* or care-giver* or guardian* or parent or parents or parental or mother* or maternal or father* or paternal or moms or dads or family or families or grandparent*) adj8 (inform* or know* or learn* or comprehension or comprehend* or question*)).mp.
21. ((information or know*) and (mak* decision* or decision making or inform choic* or make choice* or choose or choosing)).mp.
22. (inform* or know*).ti,kf. or (inform* or know*).ab. /freq=3 23
23. or/12-22
24. Emergency Treatment/ or Emergency Medicine/ or emergency medical services/ or emergency room visits/ or exp emergency service, hospital/ or emergency services, psychiatric/ or trauma centers/ or exp Evidence-Based Emergency Medicine/ or exp Emergency Nursing/ or Emergencies/ or (emergicent* or casualty department* or ((emergenc* or ED) adj2 (room* or accident or ward or wards or unit or units or department* or physician* or doctor* or nurs* or treatment* or presentation or visit or visits or setting or patient or patients or medicine or care)) or (trauma adj1 (cent* or care))).mp.
25. 5 and 8 and 11 and 23 and 24

